# Effect of cyclic daytime versus continuous enteral nutrition on circadian rhythms in critical illness: a randomized controlled trial

**DOI:** 10.64898/2026.08.24.26361187

**Authors:** Floor W. Hiemstra, Marit F. van Gent, Johanna H. Meijer, Hassan S. Dashti, Evert de Jonge, David J. van Westerloo, Laura Kervezee

**Affiliations:** Department of Intensive Care, Leiden University Medical Center, Albinusdreef 2, 2333 ZA, Leiden, The Netherlands; Group of Circadian Medicine, Department of Cell and Chemical Biology, Leiden University Medical Center, Albinusdreef 2, 2333 ZA, Leiden, The Netherlands; Department of Anesthesiology, Mass General Brigham, 55 Fruit Street, Boston, MA 02114, USA; Division of Sleep Medicine, Harvard Medical School, 25 Shattuck Street, Boston, MA 02115, USA; Division of Nutrition, Harvard Medical School, 25 Shattuck Street, Boston, MA 02115, USA

**Keywords:** Circadian Rhythm, Critical Care, Enteral Nutrition, Intensive Care Unit

## Abstract

**Objective:** Circadian rhythms are frequently disrupted in patients in the intensive care unit (ICU), potentially worsening clinical outcomes. Continuous enteral nutrition throughout the day and night is common in the ICU, but eliminates feeding-fasting cycles that serve as important timing cues for the circadian system. The objective of this study was to determine the effect of providing enteral nutrition in a cyclic daytime pattern, compared with continuous administration, on circadian rhythmicity in critically ill patients in the ICU.

**Design:** Single-center randomized controlled trial

**Setting:** Mixed medical-surgical tertiary intensive care unit in the Netherlands

**Patients:** Adult ICU patients (≥18 yr) receiving enteral nutrition.

**Intervention:** Patients were randomized to receive either continuous, or cyclic daytime enteral feeding (08:00-20:00), initiated from the start of nutritional support.

**Measurements and Main Results:** Sixty-two ICU patients were enrolled, of whom 51 were included in the per-protocol analysis. While the amplitude of the 24-hour rhythm in core body temperature did not differ significantly between the cyclic daytime and continuous feeding groups (0.17 [interquartile range: 0.09-0.24] vs. 0.20 [0.13-0.30], p=0.182), the 24-hour rhythm in heart rate was enhanced in patients receiving cyclic daytime feeding, as reflected by significantly higher amplitudes and more synchronized peak times. No significant differences in 24-hour rhythmicity were observed between groups for the other vital signs or melatonin.

**Conclusions:** Our findings suggest that cyclic daytime feeding may strengthen circadian rhythms in critically ill patients. Further studies are warranted to evaluate its impact on clinical outcomes.

**Trial registration:** Registered at ClinicalTrials.gov (number: NCT05795881).

**Key points:** *Question:* Does cyclic daytime feeding – restricting enteral nutrition to daytime hours - enhance circadian rhythmicity in vital signs in critically ill patients, compared to standard continuous enteral nutrition?

*Findings:* In this randomized controlled trial of 62 ICU patients, the amplitude of the 24-hour in core body temperature did not differ significantly between the cyclic daytime and continuous feeding groups. However, the 24-hour rhythm in heart rate was enhanced in patients receiving cyclic daytime feeding, as reflected by significantly higher amplitudes and more synchronized peak times. No significant differences in 24-hour rhythmicity were observed between groups for the other vital signs or melatonin.

*Meaning:* Our findings suggest that cyclic daytime feeding holds promise as a practical intervention to strengthen circadian rhythms in ICU patients, although further research is needed to determine whether it may affect other circadian outcomes or clinical outcomes.

## Introduction

Consisting of a central pacemaker in the hypothalamic suprachiasmatic nucleus and peripheral clocks present in nearly all body tissues, the circadian system regulates 24-hour rhythms in numerous physiological processes and thereby supports health and wellbeing. In critically ill patients in the intensive care unit (ICU), these rhythms are often profoundly disrupted (1, 2), manifesting as the absence of rhythmicity in melatonin secretion (3) and clock gene expression (4), as well as abolished, dampened or phase-shifted rhythms in heart rate (HR), blood pressure (BP) and core body temperature (CBT) (5–7). Disruption of circadian rhythms has been shown to negatively impact immune, metabolic, cardiovascular and cognitive function in non-hospitalized individuals (8–13), and has been associated with inflammation, delirium, mortality and a longer length of stay in patients in the ICU (5, 6, 14–16).

In addition to the light-dark cycle, the feeding-fasting cycle acts as a potent synchronizer for the circadian system (17). From a circadian perspective, the common ICU practice of providing enteral nutrition continuously over the 24-hour period is profoundly different from natural feeding-fasting cycles and may exacerbate circadian disruption by eliminating a potent synchronizing cue for the circadian system. Supporting this, experimental studies in animals and non-hospitalized human participants have shown that rhythmic feeding can effectively entrain peripheral clocks and restore dampened circadian patterns in metabolic and hormonal processes (18, 19) as well as in vital signs such as BP (20, 21) and CBT (22). In addition, daytime-restricted parenteral feeding was shown to result in earlier oral food intake in pediatric patients following stem cell transplantation (23). However, whether restricting enteral nutrition to the daytime may help to restore circadian rhythms in critically ill patients in the ICU remains largely unexplored (24, 25).

To address this question, we conducted a randomized controlled trial to investigate whether cyclic daytime feeding – restricting enteral nutrition to daytime hours – improves circadian rhythms in vital signs in critically ill patients compared with standard continuous enteral feeding, and to explore effects on glycemic control, nutritional delivery and tolerance, and clinical endpoints.

## Methods

### Trial design, setting and approval

The CIRCLES trial was an investigator-initiated, single center, open-label randomized controlled trial conducted between June 2023 and March 2025 at the adult ICU of Leiden University Medical Center (LUMC, Leiden, the Netherlands), a tertiary care mixed medical-surgical 38-bed ICU. The trial protocol was prospectively registered at ClinicalTrials.gov (NCT05795881; date of registration: 3 April 2023) and has been published (26). The trial is reported in accordance with the Consolidated Standards of Reporting Trials (CONSORT) guidelines for randomized controlled trials (27). Ethical approval for the trial has been granted by the local medical ethics committee of Leiden, Den Haag, and Delft in the Netherlands (P22.080; date of approval: 9 December 2022). Informed consent from patients was obtained in accordance with national regulations and the protocol reviewed by the ethics committee (see details in **eMethods 1**).

### Participants

All patients admitted to the ICU were screened for enrolment upon ICU admission, no later than the day of initiation of enteral nutrition. Inclusion criteria were: age ≥ 18 years old; receiving or intention to start enteral nutrition via nasogastric or nasoduodenal tube; expected duration of ICU admission at time of screening > 48 hours; arterial catheter in situ. The full list of exclusion criteria are available in **eMethods 2**.

### Intervention

Patients were randomly assigned to either the *continuous* enteral feeding group (control) or the *cyclic daytime* enteral feeding group (intervention) (see details in ref. (26)). In the continuous group, enteral nutrition was provided over 24 hours per day. In the cyclic daytime group, nutrition was restricted to a 12-hour period between 08:00h–20:00h. The allocated feeding protocol was followed until ICU discharge or until enteral nutrition was discontinued due to standard clinical reasons or if any of the criteria listed in **e-Methods 3** were met, after which patients received standard ICU care.

### Enteral nutrition

According to the standard clinical protocol, enteral feeding was initiated within 24 hours of ICU admission, if clinically feasible. In both groups, enteral feeding was started at a continuous rate (20 mL/h in the continuous group vs. 40 mL/h in the cyclic daytime group), with gradual increases of 20 mL/h and 40 mL/h, respectively, every 4-6 hours if gastric residual volume (GRV) remained below 200 mL, until the target was reached; see ref. (26) for details). Monitoring and management of GRV as well as treatment of feeding intolerance was similar across groups, as previously described (26).

### Glycemic control

In both study groups, blood glucose concentration was measured at least every 6 hours as part of routine clinical care, with additional checks if clinically indicated. In the cyclic daytime group, an additional daily glucose check was done 1 hour after pausing the enteral nutrition for the overnight fast (see ref. (26) for details).

### Vital signs data acquisition and processing

Vital sign data (CBT, HR, mean BP (BPm), and electrocardiogram (ECG) were automatically logged as part of routine clinical care and extracted for the purpose of the trial. Numeric data were preprocessed by removing artifacts and downsampled to 15-minute medians. HRV was derived from ECG R–R intervals and quantified as root mean square successive differences (RMSSD), calculated in 5-minute windows every 15 minutes. A detailed description of data acquisition and preprocessing is provided in **e-Methods 4**.

### Plasma melatonin sampling and analysis

To assess circadian rhythms in melatonin, arterial blood samples were collected for plasma melatonin analysis every 4 hours over a 24-h period, from 08:00h on study day 3 to 08:00h on study day 4, resulting in seven samples per patient (see **e-Methods 5** for details).

### Study outcomes

The primary outcome was the amplitude of the 24-h rhythm of CBT, assessed by cosinor analysis at study days 3-4. Secondary outcomes include measures related to 24-h rhythms in BPm, HR, HRV and melatonin, as well as glycemic control, nutritional timing, delivery and tolerance, and clinical endpoints (28-day mortality, ICU and hospital length of stay, duration of invasive mechanical ventilation, incidence of delirium and infections from the start of the study intervention until ICU discharge). No interim analysis were performed. Full details on outcomes are available in **e-Methods 6**.

### Statistics

The planned sample size was 30 patients per group who completed follow-up to study day 4 at 08:00h (see ref. (26)). Between-group comparisons were performed using Student’s t-tests, Mann-Whitney U tests, or Fisher’s exact tests, as appropriate. Cosinor acrophases and melatonin midpoints were presented as circular mean and standard deviation (SD) given the circular nature of the data, and their phase clustering was assessed using the Rayleigh test. If data were significantly clustered within groups, group differences in mean phase were tested using the Watson–Williams test. Melatonin concentrations between groups and over time were compared using a linear mixed effects model, with sampling time point and randomization group as fixed effects, along with their interaction term, and patient as random effect. No interim analyses were performed. Statistical analyses were performed in Python using the SciPy (version 1.16.0) (28), Astropy (version 7.1.0) (29), and lifelines (version 0.30.0) (30) libraries, except for linear mixed-effects modelling, which was performed in R using the lme4 package (version 1.1.37) (31).

Outcomes related to 24-hour rhythms were analyzed in the follow-up cohort (i.e., patients who completed follow-up to study day 4 at 08:00h) by both per-protocol and intention-to-treat analysis. Nutritional timing, delivery and tolerance, glycemic control and clinical endpoints were analyzed in the full cohort (i.e., all included patients) using intention-to-treat analysis. For the definition of protocol compliance, see **e-Methods 7**. Only complete study days were included in the analysis of nutritional timing, delivery and tolerance and glycemic control, excluding the day of enrolment and day of termination of the study protocol.

## Results

### Participant flow and baseline characteristics

Between June 2023 and March 2025, a total of 3,228 patients were screened for eligibility. Of these, 147 patients were randomized and consent was obtained for 85 patients (*full cohort;* **Figure 1**). Of these, 76 had at least one complete study day (42 in the continuous group and 34 in the cyclic daytime group) and were included in the analysis of the nutritional and glycemic measures, while 62 patients completed follow-up until study day 4 (*follow-up intention-to-treat cohort)*.

**Figure 1.**
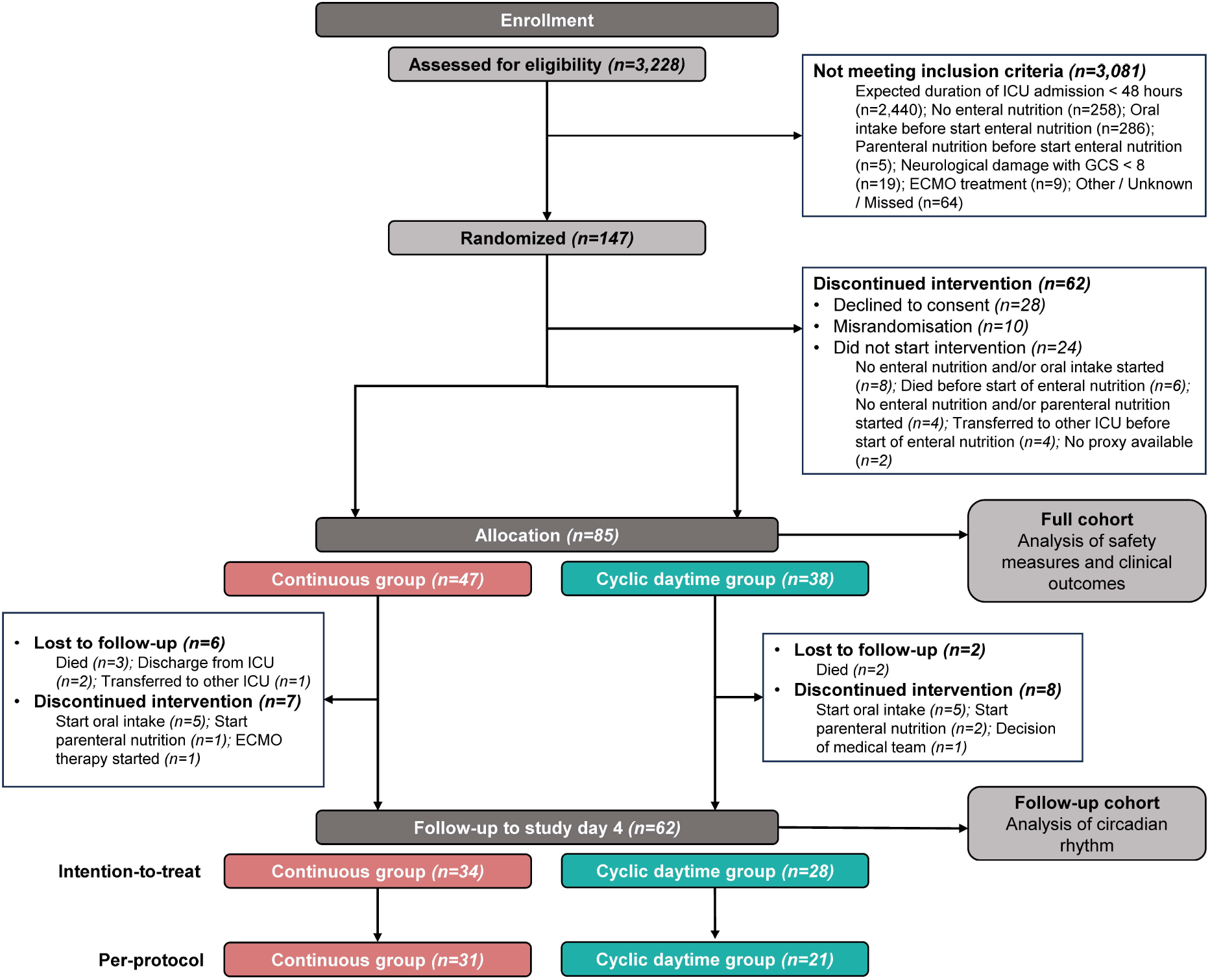
Inclusion flowchart.

Nutritional timing was assessed to determine protocol compliance and to establish the *follow-up per-protocol cohort*. Feeding times across the study of two representative patients in each group are shown in **Figure 2A and B**. As intended by protocol, feeding hours were distributed evenly across 24 hours in the continuous group (**Figure 2C**), and half of all feeding hours occurred during daytime hours (08:00h-20:00h) (50% [50-50]) (**e-Figure 1A**). Feeding hours were concentrated in daytime hours in the cyclic daytime group (**Figure 2D**), with 98% [94-100] of feeding hours delivered between 08:00h and 20:00h (**e-Figure 1B**). In total, 52 patients met the criteria for protocol compliance and were included in the per-protocol analysis (31 in the continuous group and 21 in the cyclic daytime group, *follow-up per-protocol cohort*). Baseline characteristics and patient characteristics on study days 3 and 4 were comparable between groups (**Table 1 and e-Tables 1 and 2**).

**Figure 2.**
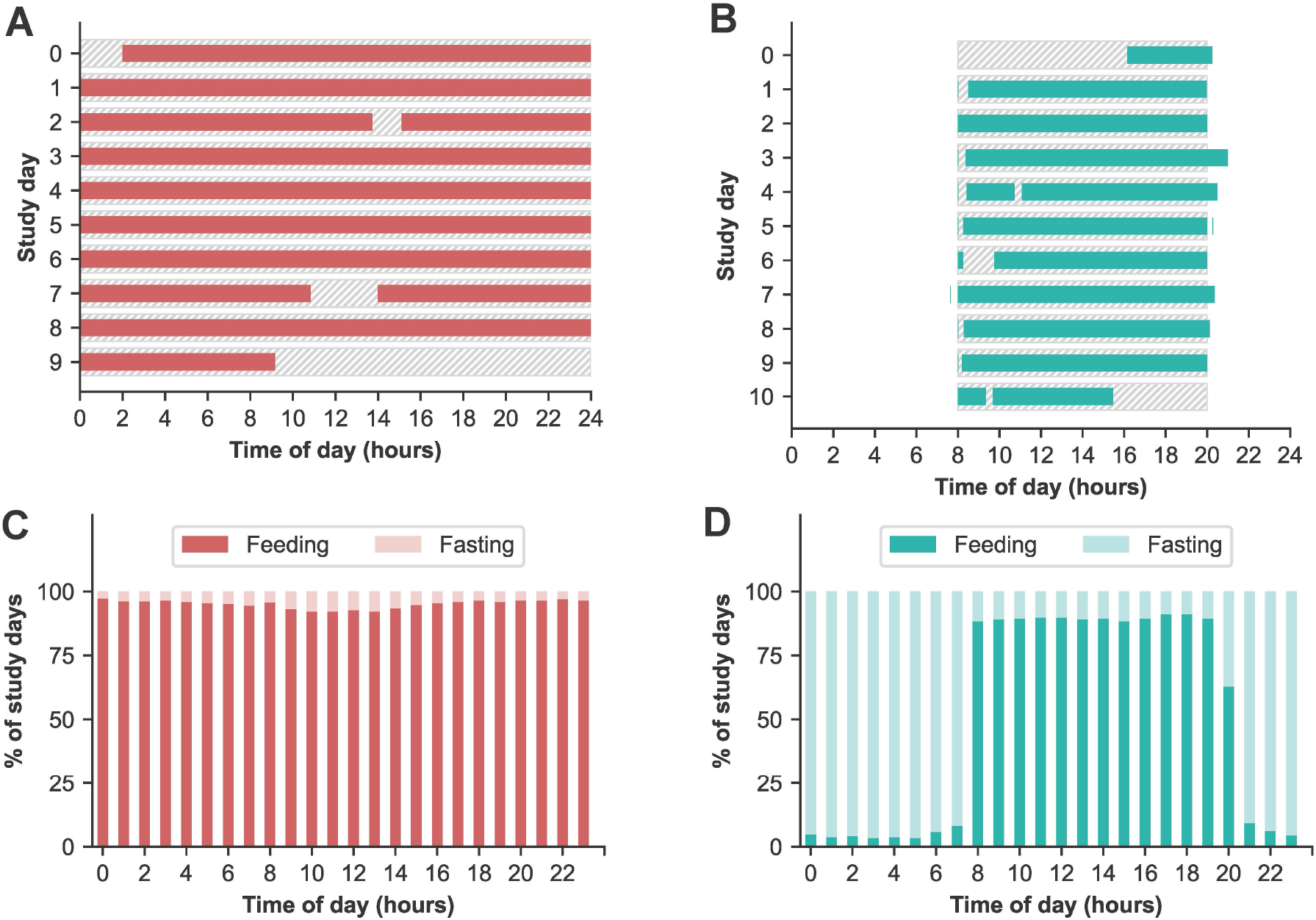
Nutritional timing and delivery (full cohort, intention-to-treat analysis). **(A-B)** Feeding times of a representative patient from the **(A)** continuous group and **(B)** cyclic daytime group. Dashed bars indicate prescribed feeding hours, filled bars indicate actual feeding hours. **(C-D)** Percentage of feeding and fasting per hour of day across all study days for the **(C)** continuous group (n=42) and **(D)** cyclic daytime group (n=34).

**Table 1.** Baseline characteristics of study population (follow-up, per-protocol cohort).

|  | Continuous group (n=31) | Cyclic daytime group (n=21) |
| --- | --- | --- |
| **Age (years), median [IQR]** | 61.0 [54.0 - 68.5] | 56.0 [48.0 - 68.0] |
| **Female sex (assigned at birth), n (%)** | 7 (23%) | 6 (29%) |
| **BMI, median [IQR]** | 26.9 [24.3 - 29.7] | 27.8 [23.9 - 31.4] |
| **APACHE IV-score, mean ± SD** | 74.8 ± 28.1 | 70.3 ± 18.3 |
| **SAPS III score, mean ± SD** | 43.6 ± 9.6 | 42.0 ± 10.3 |
| **Diabetes diagnosis, n (%)** | 7 (23%) | 4 (19%) |
| **Type of admission, n (%)** |  |  |
| Medical | 16 (51.6%) | 12 (57.1%) |
| Elective surgery | 8 (25.8%) | 4 (19.0%) |
| Emergency surgery | 7 (22.6%) | 5 (23.8%) |
| **Reason for admission, n (%)** |  |  |
| Cardiovascular | 13 (41.9%) | 10 (47.6%) |
| Respiratory | 12 (38.7%) | 3 (14.3%) |
| Transplant | 3 (9.7%) | 2 (9.5%) |
| Neurological | 2 (6.5%) | 2 (9.5%) |
| Trauma | 1 (3.2%) | 1 (4.8%) |
| Genitourinary | 0 (0%) | 1 (4.8%) |
| Metabolism | 0 (0%) | 1 (4.8%) |
| Musculoskeletal/skin | 0 (0%) | 1 (4.8%) |
| **Time from ICU admission to initiation of enteral nutrition (hours), median [IQR]** | 22.1 [14.7 - 37.5] | 19.2 [15.7 - 27.5] |
| **Type of mechanical ventilation at start of intervention, n (%)** |  |  |
| Invasive | 29 (93.5%) | 20 (95.2%) |
| Non-invasive | 2 (6.5%) | 0 (0%) |
| None | 0 (0%) | 1 (4.8%) |
| **Number of study days per patients, median [IQR]** | 7.0 [4.0-12.0] | 7.5 [4.2-15.8] |

Normally distributed continuous variables are presented as mean ± SD, non-normally distributed variables as median [IQR25–IQR75], and categorical data as n (%).

**Table 2.** Outcomes on nutritional timing, delivery and tolerance, glycemic control and clinical endpoints (full cohort, intention-to-treat analysis).

|  | Continuous group | Cyclic daytime group | p-value |
| --- | --- | --- | --- |
| **Nutritional delivery and tolerance** |  |  |  |
|  | n=42 patients<br>n=396 days | n=34 patients<br>n=293 days |  |
| Caloric intake (kcal/kg/day), mean ± SD | 24.8 ± 5.8 | 20.7 ± 7.3 | 0.007** |
| Time to reach 80% of caloric target (hours), mean (95% CI)<sup>a</sup> | 30.4 (25.3-36.1) | 63.0 (44.7-74.0) | <0.001*** |
| Target rate (kcal/day), median [IQR] | 2071 [1804-2216] | 2022 [1750-2139] | 0.530 |
| Days with high GRV (> 200 mL), n (%) | 40 (10.1%) | 50 (17.1%) | 0.008** |
| Patients with prokinetic treatment, n (%) | 25 (59.5%) | 25 (73.5%) | 0.232 |
| **Glycemic control** |  |  |  |
|  | n=42 patients<br>n=396 days | n=34 patients<br>n=293 days |  |
| Time-weighted glucose average (mmol/L), mean ± SD | 8.51 ± 1.01 | 9.21 ± 1.59 | 0.023* |
| Coefficient of variation of glucose levels (%), median [IQR] | 12.3 [8.6 - 15.1] | 18.1 [12.9 - 26.1] | <0.001*** |
| Patients with ≥ 1 hyperglycemic episode, n (%) | 35 (83.3%) | 29 (85.3%) | 1.000 |
| Patients with ≥ 1 hypoglycemic episode, n (%) | 2 (4.8%) | 3 (8.8%) | 0.651 |
| Days with ≥ 1 hyperglycemic episodes, n (%) | 188 (47.5%) | 218 (74.4%) | <0.001*** |
| Days with ≥ 1 hypoglycemic episodes, n (%) | 4 (1.0%) | 5 (1.7%) | 0.506 |
| Patients with insulin therapy, n (%) | 28 (66.7%) | 34 (100.0%) | <0.001*** |
| Days with insulin therapy, n (%) | 222 (56.1%) | 160 (54.6%) | 0.757 |
| Insulin units per day (IU/day), median [IQR] | 50.78 [21.22 - 74.25] | 38.46 [30.44 - 61.00] | 0.421 |
| **Clinical endpoints** |  |  |  |
|  | n=47 | n=38 |  |
| 28-day mortality, n (%) | 11 (23.4%) | 12 (31.6%) | 0.466 |
| ICU length of stay (days), median [IQR] | 9.0 [6.0 - 16.0] | 11.5 [6.0 - 20.5] | 0.348 |
| Hospital length of stay (days), median [IQR] | 18.0 [13.0 - 37.5] | 18.0 [12.2 - 41.0] | 0.933 |
| Duration of mechanical ventilations (days), median [IQR] | 6.00 [4.00 - 12.25] | 9.50 [4.00 - 15.00] | 0.617 |
| Incidence of delirium, n (%) | 14 (29.8%) | 11 (28.9%) | 1.000 |
| Incidence of infections, n (%) | 25 (53.2%) | 20 (52.6%) | 1.000 |

Normally distributed continuous variables are presented as mean ± SD and compared between groups using a t-test, non-normally distributed variables are presented as median [IQR25–IQR75] and compared using the Mann–Whitney U test.
<sup>a</sup>: Time to reach caloric target was compared between groups using Kaplan-Meier survival analysis and a log-rank test.
Significant differences between groups are indicated as: p<0.05 (\*), p<0.01 (\*\*), and p<0.001 (\*\*\*).
Details on infection types and sites are presented in **e-Table 8**.

### 24-hour rhythms in vital signs

**Figure 3A&B** show representative examples of patient-level cosinor models fitted to 48-hour vital sign recordings (see **e-Figure 2** for details regarding data availability per vital sign). Although cosinor amplitudes across different vital signs were consistently higher in the cyclic daytime group than in the continuous group (**Figure 3C-F**), no clear statistical evidence was found for a difference in the pre-defined primary outcome (amplitudes of the 24-hour rhythm in core body temperature) between the cyclic daytime group (median [IQR]: 0.20 [0.13 – 0.30] °C) and the continuous group (0.17 [0.09 – 0.24] °C, p=0.182, Mann-Whitney U test) (**e- Table 3**, **Figure 3C**). For the 24-hour rhythm in heart rate, amplitudes were significantly higher in patients receiving cyclic daytime feeding (**e- Table 3**; **Figure 3D**). No significant improvements in 24-hour rhythmicity were observed for blood pressure and heart rate variability (**e- Table 3**, **Figure 3E-F**). Significant phase clustering of cosinor acrophase (peak time) was observed in the cyclic daytime group in heart rate, indicating synchronization of heart rate peak times, but not for the other vital signs (**e- Table 3**; **Figure 3G-J**). Likewise, group-level analyses showed higher amplitudes in the cyclic daytime group for all vital signs (**e- Figure 3 ; e- Table 4**). The intention-to-treat analyses yielded results consistent with the primary (per-protocol) analyses (**e- Table 5).**

**Figure 3.**
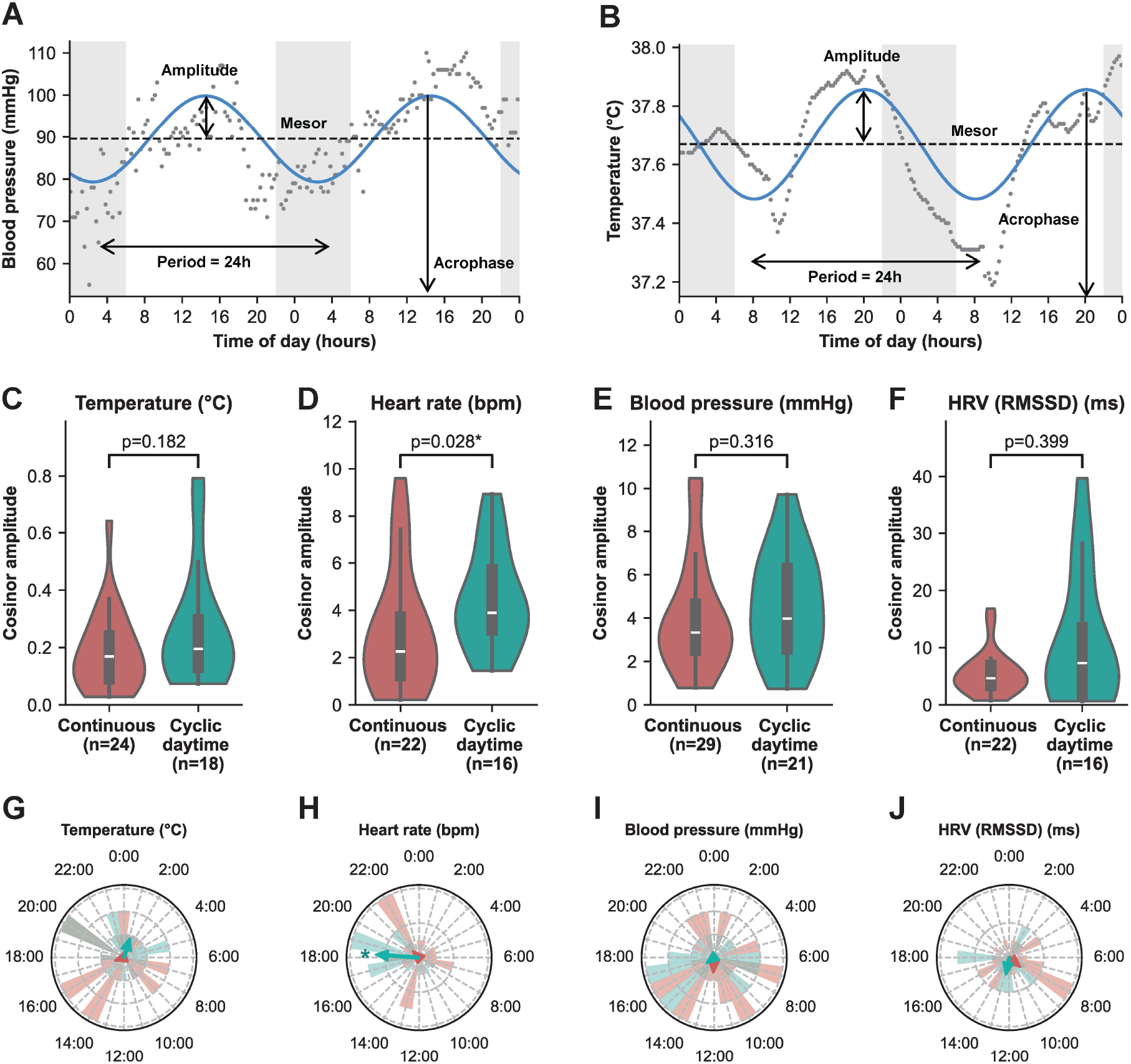
Cosinor parameters of vital signs (follow-up cohort, per-protocol analysis). **(A-B)** Representative example of cosinor fit of BPm **(A)** and CBT **(B)** over 48 hours with observed values (dark gray dots) and cosinor fit (blue line). Cosinor metrics (mesor, amplitude, acrophase and period) are indicated in black. Grey shading indicate nighttime periods (22:00h- 6:00h). **(C-F)** Cosinor amplitude for **(C)** CBT, **(D)** HR, **(E)** BPm, and **(F)** HRV. Data is shown as violin plot with the kernel density estimate of the distribution combined with a box-and- whisker plot, indicating the median and interquartile range, with the whiskers extending to 1.5 x the interquartile range. **(G-J)** Cosinor acrophase (timing of the peak) shown in polar distribution plots for **(G)** CBT, **(H)** HR, **(I)** BPm, and **(J)** HRV. The circular mean acrophase is indicated by arrows, with arrow length proportional to the resultant vector length. Significant phase clustering, as determined with Rayleigh test, is indicated with *. The continuous group is represented in red, and the cyclic daytime feeding group in teal. Sample sizes per group for each vital sign are shown in panels C-F.

### 24-hour rhythms in melatonin

Group-level analysis revealed a significant main effect of sampling time on log-transformed melatonin levels (p<0.001, linear mixed effects model), but no significant effect of group (p=0.448), or interaction between sampling time and group (p=0.963), reflecting 24-hour variation in melatonin concentrations in both groups with higher levels at night than during the day (**Figure 4A**; see **e- Figure 1** for details on data availability).

**Figure 4.**
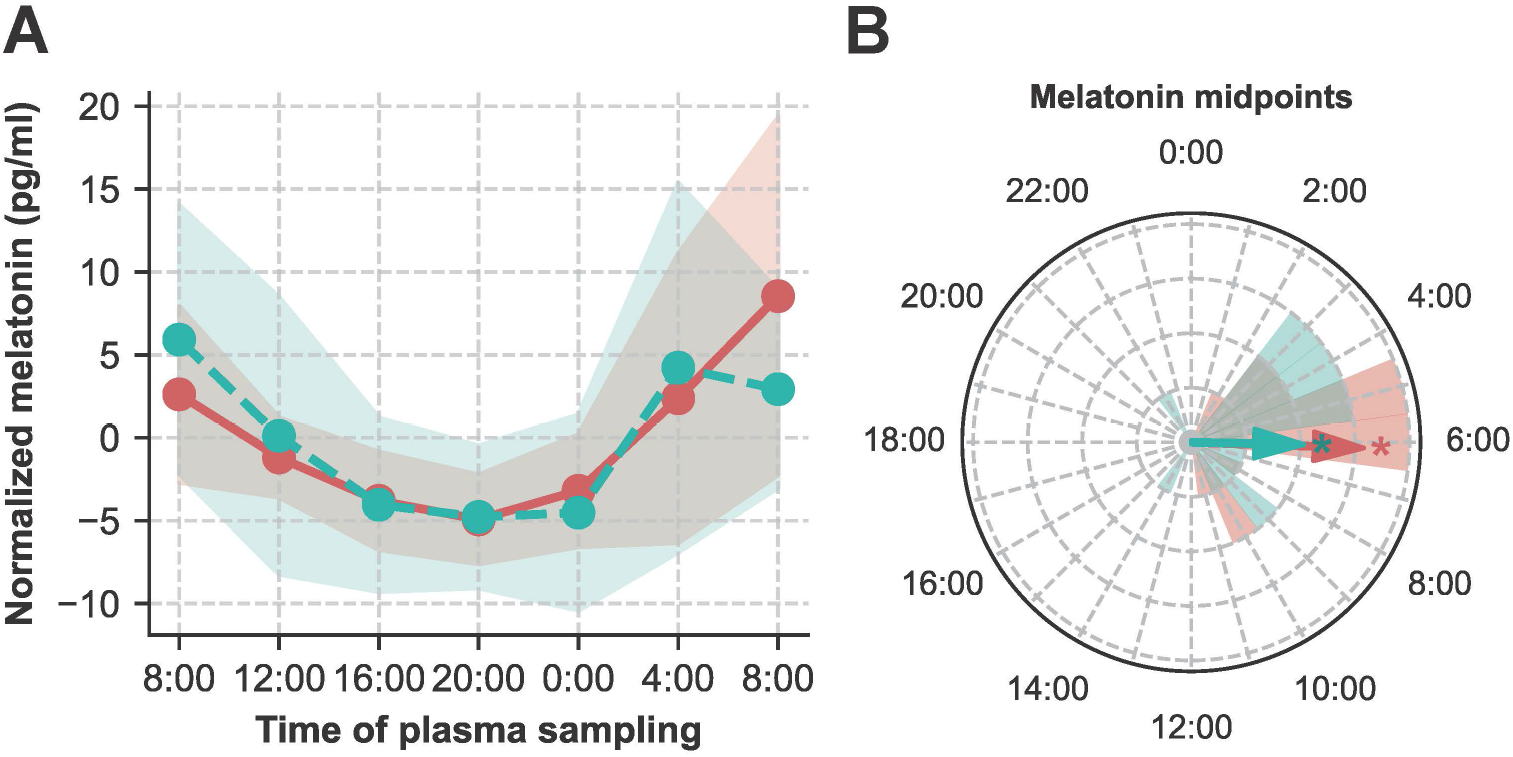
24-hour variation in plasma melatonin (follow-up cohort, per-protocol analysis). **(A)** Normalized plasma melatonin concentrations over time for continuous group (n=24, red color) and cyclic daytime group (n=17, teal color). No significant group differences (p=0.448) or sampling time × group interactions (p=0.963) were observed (linear mixed effects model), although melatonin levels varied significantly across time points (p < 0.001). Data are shown as median, with the filled area representing the interquartile range. Melatonin data was normalized by mean subtraction for each individual for visualization purposes. **(B)** Polar histogram showing the distribution of melatonin midpoints for the continuous group (n=24, red color) and cyclic daytime group (n=17, teal color). The colored bars represent the frequency of melatonin midpoints at different times of day, with the arrows indicating the circular mean and their length proportional to the resultant vector length. Significant phase clustering, as determined with Rayleigh test, is indicated with *.

Individual melatonin midpoints, which could be identified in 28 patients (**e- Table 6)**, did not differ between groups (continuous group: 06:07 ± 02:42 (hh:mm); cyclic daytime group: 05:47 ± 03:41; p=0.800, Watson–Williams test, **Figure 4B**). Similar results were obtained with intention-to-treat analysis (**e- Table 7**).

### Nutritional, glycemic, and clinical outcomes

The mean caloric intake per kilogram body weight per day across the entire study period was significantly lower in the cyclic daytime group than in the continuous group (**Table 2**). Although the proportion of patient days with a high GRV (> 200 mL) was higher in the cyclic daytime group, the proportion of patients receiving prokinetics did not differ between groups (**Table 2**). Patients in the cyclic daytime group had a small but significantly higher time-weighted glucose average and greater coefficient of variation of glucose levels than those in the continuous group, while the proportion of patients that had a hypoglycemic and hyperglycemic event in the study period did not significantly differ between groups (**Table 2**). However, on a patient-day level, hyperglycemic events occurred more frequently in the cyclic daytime group (**Table 2**). Significantly more patients received insulin therapy in the cyclic daytime group compared to the continuous group, but there was no difference in the proportion of days with insulin therapy in patients who received insulin, and on those days with insulin therapy, insulin doses were similar between groups (**Table 2**). No significant differences were found between feeding groups in exploratory clinical endpoints (28-day mortality, ICU and hospital length of stay, duration of mechanical ventilation and incidence of delirium and infections, **Table 2**).

## Discussion

In this randomized controlled trial, we investigated whether cyclic daytime feeding in the ICU supports circadian rhythmicity in vital signs in critically ill patients compared with standard continuous enteral feeding. The effect of cyclic daytime feeding was most evident on the 24- hour rhythm in HR, which showed significantly higher cosinor amplitudes and synchronized peak times across patients receiving cyclic daytime feeding. We used a pragmatic intervention that allowed for implementation within routine ICU workflows, as evidenced by good adherence to the protocol-prescribed feeding times. These findings provide evidence that cyclic daytime feeding is a feasible chronobiological intervention for strengthening circadian rhythms in critically ill patients.

Contrary to our hypothesis that cyclic daytime feeding would have most pronounced effects on the 24-hour rhythm in CBT (*a priori* defined as our primary outcome), we did not find clear evidence for this: while the values were numerically higher in patients receiving daytime feeding than in patients receiving continuous feeding, this difference was not statistically significant. The impact of the timing of nutrition on the entrainment of the endogenous circadian rhythm in CBT has been shown in healthy individuals under highly controlled conditions (32). Additionally, feeding may influence CBT through a direct thermogenic effect on core body temperature, resulting from metabolically-induced heat production. The resulting physiological fluctuation in CBT induced by daytime nutrition may, in turn, entrain other peripheral circadian rhythms (33–35). The absence of a clear effect of our intervention on CBT amplitudes may be due to the high degree of heterogeneity of the patient population and the relatively small sample size: although the study reached the pre-specified sample size, missing vital sign recordings and patients not meeting the per-protocol criteria reduced the final number of patients included in the analysis.

The effects of cyclic daytime feeding were most notable on the 24-hour rhythm of HR, with significantly higher amplitudes and synchronized and properly-phased rhythms with cyclic daytime feeding. Mechanistically, this could be due to a direct effect of nutrition on HR, an endogenous circadian effect, or a combination of both. In healthy populations, HR is known to increase acutely following food intake, contributing to a postprandial increase in cardiac output (36). In an animal model, timed feeding was shown to synchronize the peak timing and amplitude of the 24-hour rhythm in HR (and CBT) with altered feeding times (37). Whether these feeding-related physiological responses may translate into improved circadian synchronization of other physiological processes, remain to be determined in future studies.

We did not observe an effect of the intervention on melatonin rhythmicity, in line with the hypothesis that nutrition primarily acts as a potent timing cue for peripheral clocks rather than the central clock (17). However, a noteworthy observation is the majority of melatonin peak times were delayed by nearly three hours in our study population compared to healthy populations (38). Similarly, HR peaks occurred approximately two hours later in our study than in a cohort of age-matched, non-hospitalized adults (39). These phase delays are consistent with previous reports in critically ill populations, with phase delays reported in 24-hour rhythm of HR(40), CBT(7), melatonin, as well as blood glucose levels (41–43). The circadian phase delays imply that some patients in our study may have received enteral nutrition in part during their biological night, when melatonin levels were still elevated. This may have metabolic relevance, as feeding during the biological night is known to impair glucose regulation (8, 44). An important consideration for future research is therefore to ensure that feeding schedules are aligned with patients’ circadian phase.

From a clinical perspective, we observed minor differences between groups in nutritional delivery and glycemic control, but no severe adverse events were reported, and the intervention was generally well tolerated. The lower caloric intake observed in the cyclic daytime group may be explained by the relatively low GRV threshold of 200 mL used to titrate infusion rates in our ICU at the time of the study, which may have delayed the achievement of nutritional targets. However, routine GRV monitoring is considered a poor marker of gastrointestinal tolerance and is no longer recommended in current ESPEN critical care guidelines (45). In future studies, initiating cyclic daytime feeding after nutritional targets are achieved may better balance adequate caloric intake with potential circadian benefits (46). Furthermore, we observed slightly but significantly higher glucose levels in the cyclic daytime feeding group, possibly explained by higher daytime feeding rates. Despite this, daily insulin use did not differ between groups, possibly due to clinicians’ hesitation to use higher insulin rates during the higher daytime nutritional delivery rates in the cyclic feeding group. Thus, these findings likely reflect implementation-related factors rather than an inherent fundamental disadvantage of circadian-aligned feeding.

Several limitations should be acknowledged. Firstly, we assessed the effect of the intervention on circadian rhythms relatively early in the ICU stay, when the majority of patients were still experiencing acute critical illness and the intervention may not have exerted its full effect. Future studies should explore the optimal timing and duration of cyclic daytime feeding. Moreover, it should be noted that various factors, including medications, the pathophysiology of the critical illness, inflammatory processes, and the ICU environment can interfere with the circadian biomarkers used. As a result, it is challenging to determine to what extent the observed 24-hour rhythms truly reflect endogenous circadian rhythms.

## Conclusion

In conclusion, our findings suggest that cyclic daytime feeding shows promise as a practical intervention to strengthen circadian rhythms in ICU patients, with the most profound impact on the 24-hour rhythm in HR. The feeding intervention was safe, pragmatic, and well-implemented, which opens up the opportunity to integrate timing of feeding as one component of a broader circadian-informed ICU care strategy. Follow-up studies are warranted to determine whether this intervention can translate into clinical meaningful improvements in both short- and long-term patient outcomes.

## Data Availability

Deidentified individual participant data that underlie the results reported in this article and the statistical code are available from the corresponding author upon reasonable request, subject to approval and a data sharing agreement.

## Acknowledgements

We would like to thank the ICU staff for their support in conducting the study, Rene Sterk for his assistance with the data export, Delilah van Swieten for her assistance with study coordination and data collection, and Mike del Prado and Jeanette Wigbers for their support in coordinating the study.

## Author contribution

FWH: data curation, Formal analysis, investigation, project administration, validation, visualization, writing – original draft

MFvG: data curation, investigation, writing – review & editing

JHM: writing – review & editing

HSD: writing – review & editing EdJ: writing – review & editing

DJvW: conceptualization, funding acquisition, investigation, supervision, writing – review & editing

LK: conceptualization, funding acquisition, investigation, supervision, writing – review & editing

## References

1. Knauert MP, Ayas NT, Bosma KJ, et al: Causes, Consequences, and Treatments of Sleep and Circadian Disruption in the ICU: An Official American Thoracic Society Research Statement. Am J Respir Crit Care Med 2023; 207(7):e49–e68

2. Felten M, Dame C, Lachmann G, et al: Circadian rhythm disruption in critically ill patients. Acta Physiol (Oxf) 2023; 238(1):e13962

3. Olofsson K, Alling C, Lundberg D, et al: Abolished circadian rhythm of melatonin secretion in sedated and artificially ventilated intensive care patients. Acta Anaesthesiol Scand 2004; 48(6):679–684

4. Maas MB, Iwanaszko M, Lizza BD, et al: Circadian Gene Expression Rhythms During Critical Illness. Crit Care Med 2020; 48(12):e1294–e1299

5. Davidson S, Villarroel M, Harford M, et al: Day-to-day progression of vital-sign circadian rhythms in the intensive care unit. Crit Care 2021; 25(1):156

6. Beyer SE, Salgado C, Garcao I, et al: Circadian rhythm in critically ill patients: Insights from the eICU Database. Cardiovasc Digit Health J 2021; 2(2):118–125

7. Gazendam JAC, Van Dongen HPA, Grant DA, et al: Altered circadian rhythmicity in patients in the ICU. Chest 2013; 144(2):483–489

8. Scheer FA, Hilton MF, Mantzoros CS, et al: Adverse metabolic and cardiovascular consequences of circadian misalignment. Proc Natl Acad Sci U S A 2009; 106(11):4453–4458

9. Cuesta M, Boudreau P, Dubeau-Laramee G, et al: Simulated Night Shift Disrupts Circadian Rhythms of Immune Functions in Humans. J Immunol 2016; 196(6):2466–2475

10. Morris CJ, Purvis TE, Hu K, et al: Circadian misalignment increases cardiovascular disease risk factors in humans. Proc Natl Acad Sci U S A 2016; 113(10):E1402–1411

11. Kervezee L, Kosmadopoulos A, Boivin DB: Metabolic and cardiovascular consequences of shift work: The role of circadian disruption and sleep disturbances. Eur J Neurosci 2020; 51(1):396–412

12. Fishbein AB, Knutson KL, Zee PC: Circadian disruption and human health. J Clin Invest 2021; 131(19):e148286

13. Fishbein AB, Knutson KL, Zee PC: Circadian disruption and human health. J Clin Invest 2021; 131(19)

14. Daou M, Telias I, Younes M, et al: Abnormal Sleep, Circadian Rhythm Disruption, and Delirium in the ICU: Are They Related? Front Neurol 2020; 11:549908

15. Li J, Cai S, Liu X, et al: Circadian rhythm disturbance and delirium in ICU patients: a prospective cohort study. BMC Anesthesiol 2023; 23(1):203

16. Cunningham PS, Kitchen GB, Jackson C, et al: ClinCirc identifies alterations of the circadian peripheral oscillator in critical care patients. J Clin Invest 2023; 133(4)

17. Dibner C, Schibler U, Albrecht U: The mammalian circadian timing system: organization and coordination of central and peripheral clocks. Annu Rev Physiol 2010; 72:517–549

18. Niu Y, Heddes M, Altaha B, et al: Targeting the intestinal circadian clock by meal timing ameliorates gastrointestinal inflammation. Cell Mol Immunol 2024; 21(8):842–855

19. Wehrens SMT, Christou S, Isherwood C, et al: Meal Timing Regulates the Human Circadian System. Curr Biol 2017; 27(12):1768–1775 e1763

20. Hou T, Su W, Duncan MJ, et al: Time-restricted feeding protects the blood pressure circadian rhythm in diabetic mice. Proc Natl Acad Sci U S A 2021; 118(25)

21. Zhang D, Colson JC, Jin C, et al: Timing of Food Intake Drives the Circadian Rhythm of Blood Pressure. Function 2020; 2(1)

22. Schilperoort M, van den Berg R, Dolle MET, et al: Time-restricted feeding improves adaptation to chronically alternating light-dark cycles. Sci Rep 2019; 9(1):7874

23. Wang YM, Taggart CB, Huber JF, et al: Daytime-restricted parenteral feeding is associated with earlier oral intake in children following stem cell transplant. J Clin Invest 2023; 133 (4)

24. Chiu YH, Sharma A, Dashti HS: Circadian rhythms, metabolism, and nutrition support in critically ill adult patients: a narrative review. Curr Opin Clin Nutr Metab Care 2025; 28(2):134–139

25. Dashti HS, Wang YM, Knauert MP: Feeding critically ill patients at the right time of day. Critical Care 2024; 28(1)

26. Hiemstra FW, van Gent MF, de Jonge E, et al: Effect of cyclic daytime versus continuous enteral nutrition on circadian rhythms in critical illness (CIRCLES): Study protocol for a randomized controlled trial. Contemp Clin Trials 2025:107927

27. Hopewell S, Chan AW, Collins GS, et al: CONSORT 2025 statement: updated guideline for reporting randomised trials. Lancet 2025

28. Virtanen P, Gommers R, Oliphant TE, et al: SciPy 1.0: fundamental algorithms for scientific computing in Python. Nature Methods 2020; 17(3):261–272

29. Robitaille TP, Tollerud EJ, Greenfield P, et al: Astropy: A community Python package for astronomy. Astron Astrophys 2013; 558

30. Davidson-Pilon C: lifelines: survival analysis in Python. Journal of Open Source Software 2019; 4(40):1317

31. Bates D, Machler M, Bolker BM, et al: Fitting Linear Mixed-Effects Models Using lme4. J Stat Softw 2015; 67 (1) :1–48

32. Krauchi K, Cajochen C, Werth E, et al: Alteration of internal circadian phase relationships after morning versus evening carbohydrate-rich meals in humans. J Biol Rhythms 2002; 17(4):364–376

33. Brown SA, Zumbrunn G, Fleury-Olela F, et al: Rhythms of mammalian body temperature can sustain peripheral circadian clocks. Curr Biol 2002; 12(18):1574–1583

34. Buhr ED, Yoo SH, Takahashi JS: Temperature as a universal resetting cue for mammalian circadian oscillators. Science 2010; 330(6002):379–385

35. Damiola F, Le Minh N, Preitner N, et al: Restricted feeding uncouples circadian oscillators in peripheral tissues from the central pacemaker in the suprachiasmatic nucleus. Genes Dev 2000; 14(23):2950–2961

36. Waaler BA, Eriksen M, Toska K: The Effect of Meal Size on Postprandial Increase in Cardiac-Output. Acta Physiol Scand 1991; 142(1):33–39

37. Ono M, Burgess DE, Johnson SR, et al: Feeding behavior modifies the circadian variation in RR and QT intervals by distinct mechanisms in mice. Am J Physiol Regul Integr Comp Physiol 2024; 327(1):R109–R121

38. Revell VL, Kim H, Tseng CY, et al: Circadian phase determined from melatonin profiles is reproducible after 1 wk in subjects who sleep later on weekends. J Pineal Res 2005; 39(2):195–200

39. Natarajan A, Gleichauf K, Khalid M, et al: Circadian rhythm of heart rate and activity: A cross-sectional study. Chronobiol Int 2025; 42(1):108–121

40. Knauert MP, Murphy TE, Doyle MM, et al: Pilot Observational Study to Detect Diurnal Variation and Misalignment in Heart Rate Among Critically Ill Patients. Frontiers in Neurology 2020; 11

41. Melone MA, Becker TC, Wendt LH, et al: Disruption of the circadian rhythm of melatonin: A biomarker of critical illness severity. Sleep Med 2023; 110:60–67

42. Gehlbach BK, Chapotot F, Leproult R, et al: Temporal disorganization of circadian rhythmicity and sleep-wake regulation in mechanically ventilated patients receiving continuous intravenous sedation. Sleep 2012; 35(8):1105–1114

43. Hiemstra FW, Stenvers DJ, Kalsbeek A, et al: Daily variation in blood glucose levels during continuous enteral nutrition in patients on the intensive care unit: a retrospective observational study. EBioMedicine 2024; 104:105169

44. Stenvers DJ, Scheer F, Schrauwen P, et al: Circadian clocks and insulin resistance. Nat Rev Endocrinol 2019; 15(2):75–89

45. Singer P, Blaser AR, Berger MM, et al: ESPEN guideline on clinical nutrition in the intensive care unit. Clin Nutr 2019; 38(1):48–79

46. Russell-Murray K, Dashti HS: Real-world implementation of a standardized ICU protocol for daytime-restricted enteral nutrition in critically ill adults: A retrospective quality improvement study. Clinical Nutrition Open Science 2026; 65

