## Supplementary Material for "Effect of cyclic daytime versus continuous enteral nutrition on circadian rhythms in critical illness: a randomized controlled trial"

**Short title:** Cyclic Daytime Enteral Nutrition and Circadian Rhythmicity

**Authors:** Floor W. Hiemstra MSc[1,2], Marit F. van Gent BSc[1,2], Johanna H. Meijer PhD[2], Hassan S. Dashti PhD[3,4,5], Evert de Jonge PhD[1], David J. van Westerloo PhD[1*], Laura Kervezee PhD[1,2*]

[1] Department of Intensive Care, Leiden University Medical Center, Albinusdreef 2, 2333 ZA, Leiden, The Netherlands

[2] Group of Circadian Medicine, Department of Cell and Chemical Biology, Leiden University Medical Center, Albinusdreef 2, 2333 ZA, Leiden, The Netherlands

[3] Department of Anesthesiology, Mass General Brigham, 55 Fruit Street, Boston, MA 02114, USA

[4] Division of Sleep Medicine, Harvard Medical School, 25 Shattuck Street, Boston, MA 02115, USA

[5] Division of Nutrition, Harvard Medical School, 25 Shattuck Street, Boston, MA 02115, USA

*These authors contributed equally and share corresponding authorship.

**Corresponding authors:**

L. Kervezee

Department of Intensive Care, Leiden University Medical Center

Albinusdreef 2, 2333 ZA, Leiden, The Netherlands

and

D.J. van Westerloo

Department of Intensive Care, Leiden University Medical Center

Albinusdreef 2, 2333 ZA, Leiden, The Netherlands

### Supplementary Methods

**e-Methods 1: Informed consent procedure**

Informed consent from patients was obtained in accordance with national regulations and the protocol reviewed by the ethics committee. Due to the limited time from inclusion to the start of the intervention, deferred or retrospective consent was used. If consent was not obtained within 3 days after the intervention began, patients were withdrawn from the study. Consent was either obtained directly from the patient, or – in case the patient was unable to provide consent due to their critical condition – from a proxy or legal representative. In these cases, patient consent was obtained once the patient regained decisional capacity. If a patient died before informed consent was obtained, their study data were included in the analysis.

**e-Methods 2: Exclusion criteria**

Exclusion criteria were: receiving parenteral nutrition; concurrent oral intake; prior night-time (20:00h – 08:00h) enteral or parenteral nutrition within the same hospitalization; chronic enteral tube feeding prior to current admission; presence of one or more contraindications for enteral feeding and/or at significant risk for gastrointestinal intolerance; glycemic emergency; treatment with extracorporeal membrane oxygenation (ECMO); severe neurological damage (i.e., significant neurological abnormalities such as bleeding, ischemia, neurotrauma or severe encephalopathy) with Glasgow Coma Scale (GCS) ≤ 8); suspected or confirmed pregnancy; prior study inclusion (in the case of ICU readmission).

**e-Methods 3: Criteria for discontinuation of the feeding protocol**

- Initiation of parenteral nutrition as supplement to enteral nutrition;
- oral intake as supplement to enteral nutrition exceeding 50% of total caloric needs;
- decision of treating ICU physician (e.g., due to persistent feeding intolerance despite standard of care interventions such as duodenal tube placement and prokinetic therapy);
- initiation of ECMO treatment; severe neurological damage with Glasgow Coma Scale ≤ 8 (not including sedation).

**e-Methods 4: Vital sign data acquisition and processing**

CBT, HR, mean BP (BPm) and electrocardiogram (ECG) were continuously monitored as part of routine clinical care via a patient monitor at the bedside (Philips Intellivue MP70, Philips Medical Systems, the Netherlands) and automatically logged in a data warehouse (PIIC iX, Data Warehouse Connect, Philips Medical Systems, the Netherlands). CBT was primarily measured via a bladder catheter equipped with a temperature sensor, and if unavailable, via a rectal probe. BPm was measured via an arterial line and HR was measured from the ECG system. Numeric data (CBT, HR, BPm) were stored at a sampling rate of 1 Hz, and waveform data (ECG) at 500 Hz.

All vital sign data processing was done in Python programming language.[1] Preprocessing of the numeric data included removal of values outside predefined physiological ranges (40 bpm < HR < 200 bpm, 30 mmHg < BPm < 150 mmHg, 34°C < CBT < 42°C), exclusion of segments with unphysiological fluctuations based on the time series derivative, and downsampling of the data to 15-minute intervals using the median value per 15-minute interval.

Heart rate variability (HRV) metrics were retrieved from R-R intervals derived from the continuous ECG waveform data using a modified version of the Pan-Tompkins algorithm.[2] The ECG signal was first filtered with a first-order Butterworth bandpass filter, using cutoff frequencies of 5 Hz and 15 Hz. R-peaks were then detected using the BioSPPy toolbox,[3] which differentiates and squares the signal to enhance QRS complexes for accurate R-peak detection. False-positive detections were reduced by excluding peaks with amplitudes deviating more than 1.6 times above or below one-third of the mean R-peak amplitude. Subsequently, artifacts and ectopic beats were removed from the RR interval data through the Absolute Difference between Adjacent R-R Intervals (ADARRI) method.[4] Finally, HRV was quantified using the root mean square successive differences (RMSSD) of R-R intervals - reflecting short-term, vagally mediated HRV - using the PyHRV toolbox.[5] HRV was computed in 5-minute windows every 15 minutes throughout the recording period.

For HR and HRV, values during periods with cardiac rhythm disorders or pacemaker activity were set to missing. For all vital signs, recordings containing more than 10 consecutive hours of missing data, or with less than 32 hours of valid data between the first and last data points, were excluded from analysis.

**e-Methods 5: Plasma melatonin sampling and analysis**

At each timepoint, 4 mL of blood was drawn via the indwelling arterial line, most commonly placed in the radial artery, in K2- ethylenediaminetetraacetic acid (EDTA) tubes. Blood samples were promptly processed by the hospital laboratory and centrifuged for 10 minutes at 2350g and 20°C, after which the plasma was transferred to opaque containers and stored at -80°C. Plasma melatonin concentrations were measured using liquid chromatography coupled with tandem mass spectrometry (LC-MS/MS).[6]

**e-Methods 6: Study outcomes**

*24-hour rhythm characteristics of vital signs*

All 24-hour rhythm analyses of vital signs were applied to the processed 48-hour time series covering study day 3 and 4. Amplitudes and acrophases of the 24-hour rhythms in vital signs were determined using cosinor analysis. Cosinor analysis with a fixed 24-hour period was applied to each individual time series (48 hours, covering study day 3 and 4). Cosinor analysis is a method used to model rhythmic biological data by fitting a cosine function to time series (**Figure 4A-B**). The cosinor model was fitted with generalized least squares and a first-order autoregressive error term to account for the temporal relation of the residuals using *nlme* package (version 3.1.162) in R.[7-9] Cosinor metrics (mesor, amplitude, acrophase) were extracted from the fitted cosine model. The mesor is the rhythm-adjusted mean level of the variable. The amplitude was calculated as the difference between model-fitted maximum and the mesor, indicating the strength of the 24-hour rhythm. The acrophase is the time of the peak of the fitted cosine curve. Finally, group-level analysis was performed for each vital sign by normalizing each individual's time series to their mean, calculating the median across all patients for each 15-minute interval, and fitting a cosinor model to the aggregated time series.

*24-hour rhythm characteristics of melatonin*

Patients with at least five out of seven plasma melatonin samples collected and receiving were included in the analysis and patients who received melatonin supplementation during the sampling period were excluded from the melatonin analysis. Melatonin concentrations below the detection limit (1.9 pg/mL) were imputed as 0.8 pg/mL. Melatonin midpoints and day/night ratios were derived from the individual melatonin profiles. To this end, melatonin midpoints, as a measure of central circadian phase, were determined as the time halfway between the rising and falling crossing points at 50% of the maximum melatonin level.[10] Two independent researchers (FWH and LK) visually identified the rising and falling crossing points. If no reliable crossing points were identifiable, the profile was excluded from analysis. In cases of discrepancy, consensus was reached through discussion.

*Nutritional timing, delivery and tolerance*

The feasibility of the feeding intervention was assessed in terms of nutritional timing, delivery, and tolerance. Nutritional timing was quantified as the proportion of hours when feeding was administered occurring during daytime hours. Nutritional delivery was assessed by the daily caloric intake (expressed as kcal/kg/day) from enteral nutrition and propofol administration, and the time to reach 80% of the target rate. Gastrointestinal tolerance was evaluated through the proportion of patient days with GRV exceeding 200 mL, and the proportion of patients receiving prokinetic treatment (i.e., metoclopramide, domperidone) within the study period. Measures were calculated for each study day and summarized per patient using the median across study days, where applicable. Data on nutritional timing, calories and GRV were extracted from the electronic health records.

*Glycemic control*

Glycemic control was assessed using the time-weighted average of blood glucose levels, defined as the area under the curve divided by the time between the first and last glucose check.[11] Variation in glucose levels across the study days was quantified by the coefficient of variation (standard deviation/mean x 100). Additionally, the number of patients and days with at least one hypoglycemic (blood glucose < 3.5 mmol/L), at least one hyperglycemic episode (blood glucose > 10 mmol/L) and those receiving insulin therapy were recorded. The intensity of insulin treatment was evaluated by the total number of insulin units administered per day during days with insulin therapy. Measures of glycemic control were calculated for each study day and summarized per patient using the median across study days, where applicable. Data on blood glucose levels and insulin treatment were extracted from the electronic health records.

*Clinical endpoints*

Clinical endpoints included 28-day mortality, ICU and hospital length of stay (in calendar days), duration of invasive mechanical ventilation (in calendar days), incidence of delirium and infections from the start of the study intervention until ICU discharge. Delirium was defined as a positive Confusion Assessment Method for the ICU (CAM-ICU) score,[12] or the use of haloperidol > 2 days in combination with clinical documentation of delirium. Infections were defined as a positive culture in combination with non-prophylactic use of antibiotics > 3 days. Data on clinical endpoints were extracted from the electronic health records.

**e-Methods 7: Definition of protocol compliance**

For the per-protocol analysis, protocol compliance was defined based on the proportion of feeding hours that occurred within the daytime window (08:00h-20:00h) during study days 1 to 4. In the continuous group, protocol compliance was defined as > 25% and < 75% of all feeding hours delivered during the daytime period across all study days 1 to 4 (since 50% of the feeding is expected to occur during the daytime in this group). In the cyclic daytime group protocol compliance was defined as a daytime feeding proportion of > 75% across all study days 1 to 4 (since 100% of the feeding hours is expected to occur during the daytime in this group).

### Supplementary Tables

**e-Table 1: Baseline characteristics of the full cohort (intention-to-treat).**

Abbreviations: APACHE = Acute Physiology and Chronic Health Evaluation; BMI = Body Mass Index; ICU = Intensive Care Unit; IQR = Interquartile Range; SAPS = Simplified Acute Physiology Score; SD = Standard Deviation.

| | Continuous group (n=47) | Cyclic daytime group (n=38) |
|---|---|---|
| **Age (years), median [IQR]** | 61.0 [53.5 - 68.5] | 60.5 [52.3 - 69.0] |
| **Female sex, n (%)** | 12 (26.0%) | 9 (24.0%) |
| **BMI, median [IQR]** | 27.2 [24.3 - 29.2] | 27.8 [24.2 - 31.6] |
| **APACHE IV-score, mean (± SD** | 75.7 ± 27.5 | 77.4 ± 23.4 |
| **SAPS III score, mean (± SD)** | 44.1 ± 9.5 | 44.1 ± 9.9 |
| **Diabetes diagnosis, n (%)** | 8 (17.0%) | 7 (18.4%) |
| **Type of admission, n (%)** | | |
| Medical | 27 (57.4%) | 23 (60.5%) |
| Elective surgery | 11 (23.4%) | 7 (18.4%) |
| Emergency surgery | 9 (19.1%) | 8 (21.1%) |
| **Reason for admission, n (%)** | | |
| Cardiovascular | 21 (44.7%) | 20 (52.6%) |
| Respiratory | 16 (34.0%) | 6 (15.8%) |
| Transplant | 4 (8.5%) | 4 (10.5%) |
| Neurological | 2 (4.3%) | 2 (5.3%) |
| Trauma | 1 (2.1%) | 2 (5.3%) |
| Gastrointestinal | 3 (6.4%) | 1 (2.6%) |
| Genitourinary | 0 (0%) | 1 (2.6%) |
| Metabolism | 0 (0%) | 1 (2.6%) |
| Musculoskeletal/skin | 0 (0%) | 1 (2.6%) |
| **Time from ICU admission to initiation of enteral nutrition (hours), median [IQR]** | 20.5 [14.2 - 35.4] | 21.0 [15.7 - 34.8] |
| **Type of mechanical ventilation at start of intervention, n (%)** | | |
| Invasive | 43 (91.5%) | 35 (92.1%) |
| Non-invasive | 2 (4.3%) | 2 (5.3%) |
| None | 2 (4.3%) | 1 (2.6%) |

**e-Table 2: Patient characteristics at study day 3 and 4 (follow-up, per-protocol cohort).**

[a]: Assessed at start of study day 3, [b]: RASS scores are missing in 11 patients, [c]: Assessed during study day 3 and 4. Medication classification was based on Anatomical Therapeutic Chemical (ATC) codes as follows: Analgesics: N02; Antipyretics: N02BE51 (paracetamol), N02BA01 (acetylsalicylic acid), M01AE01 (ibuprofen), M01AE52 (ibuprofen + paracetamol), M01AC06 (naproxen); Beta blockers: C07; Diuretics: C03; Glucocorticoids: H02AB; Intropics: C01CA07 (dobutamine), C01CE02 (milrinone); Melatonin (agonists): N05CH; Sedatives: N05CD08 (midazolam), N01AX10 (propofol), N05CM18 (dexmedetomidine), N05BA06 (lorazepam), C02AC01 (clonidine); Sleep medication: N05CD07 (temazepam), N05CF01 (zopiclon), N05BA04 (oxazepam); Vasopressors: H01BA04 (terlipressine), C01CA03 (norepinephrine). Abbreviations: ICU = Intensive Care Unit; IQR = Interquartile Range; RASS = Richmond Agitation-Sedation Scale; SD = Standard Deviation; SOFA = Sequential Organ Failure Assessment.

| | Continuous group (n=31) | Cyclic daytime group (n=21) |
|---|---|---|
| **Days in ICU, median [IQR][a]** | 3.4 [3.1 - 4.0] | 3.3 [3.2 - 3.7] |
| **SOFA score, mean ± SD[a]** | 9.3 ± 3.9 | 9.6 ± 3.5 |
| **RASS score, median [IQR][a,b]** | -3.0 [-4.0 - 0.0] | -3.0 [-4.0 - 0.5] |
| **Type of mechanical ventilation, n (%)[a]** | | |
| Invasive | 29 (93.5%) | 20 (95.2%) |
| Non-invasive | 2 (6.5%) | 0 (%) |
| None | 0 (%) | 1 (4.8%) |
| **Medication use, n (%)[c]** | | |
| Analgesics | 29 (93.5%) | 20 (95.2%) |
| Antipyretics | 28 (90.3%) | 20 (95.2%) |
| Beta blockers | 4 (12.9%) | 3 (14.3%) |
| Diuretics | 21 (67.7%) | 15 (71.4%) |
| Glucocorticoids | 19 (61.3%) | 12 (57.1%) |
| Inotropics | 5 (16.1%) | 7 (33.3%) |
| Melatonin (agonists) | 2 (6.5%) | 0 (0%) |
| Sedative | 19 (61.3%) | 17 (81.0%) |
| Sleep medication | 9 (29.0%) | 4 (19.0%) |
| Vasopressors (norepinephrin) | 19 (61.3%) | 14 (66.7%) |

**e-Table 3. Outcomes of cosinor and non-parametric analyses of 24-hour rhythms in vital signs (follow-up cohort, per-protocol analysis).**

| | Continuous group | Cyclic daytime group | p-value |
|---|---|---|---|
| **Core body temperature** | n=24 | n=18 | |
| Cosinor amplitude (ºC), median [IQR] | 0.17 [0.09 - 0.24] | 0.20 [0.13 - 0.30] | 0.182 |
| Cosinor acrophase (hh:mm), circular mean ± circular SD (Rayleigh test p-value) | 16:45 ± 07:38 (p=0.649) | 01:08 ± 06:01 (p=0.225) | n/a[a] |
| Cosinor mesor (ºC), mean ± SD | 37.45 ± 0.64 | 37.29 ± 0.58 | 0.405 |
| **Heart rate** | n=22 | n=16 | |
| Cosinor amplitude (bpm), median [IQR] | 2.3 [1.2 - 3.7] | 3.9 [3.2 - 5.7] | 0.028* |
| Cosinor acrophase (hh:mm), circular mean ± circular SD (Rayleigh test p-value) | 21:01 ± 07:42 (p=0.693) | 18:15 ± 03:42 (p=0.001**) | n/a[a] |
| Cosinor mesor (bpm), mean ± SD | 84.9 ± 17.0 | 82.2 ± 13.5 | 0.579 |
| **Mean blood pressure** | n=29 | n=21 | |
| Cosinor amplitude (mmHg), median [IQR] | 3.3 [2.5 - 4.7] | 4.0 [2.6 - 6.3] | 0.316 |
| Cosinor acrophase (hh:mm), circular mean ± circular SD (Rayleigh test p-value) | 12:13 ± 06:39 (p=0.254) | 15:31 ± 07:39 (p=0.689) | n/a[a] |
| Cosinor mesor (mmHg), mean ± SD | 78.7 ± 10.8 | 79.6 ± 9.9 | 0.778 |
| **Heart rate variability (RMSSD)** | n=22 | n=16 | |
| Cosinor amplitude (ms), median [IQR] | 4.65 [3.37 - 6.94] | 7.29 [1.83 - 13.54] | 0.399 |
| Cosinor acrophase (hh:mm), circular mean ± circular SD (Rayleigh test p-value) | 08:39 ± 06:46 (p=0.392) | 12:45 ± 05:40 (p=0.174) | n/a[a] |
| Cosinor mesor (ms), median [IQR] | 15.21 [11.55 - 33.39] | 22.86 [13.38 - 64.05] | 0.220 |

Normally distributed continuous variables are presented as mean ± SD and compared between groups using a t-test, non-normally distributed variables are presented as median [IQR25–IQR75] and compared using the Mann–Whitney U test.
[a]: Group comparison was not performed because significant phase clustering (Rayleigh test) was observed in none of the groups or only one group.
Significant differences between groups are indicated as: p<0.05 (*), p<0.01 (**), and p<0.001 (***).

**e-Table 4: Cosinor parameters of group-level fits (follow-up cohort, per-protocol analysis).**

Data was normalized by mean subtraction per individual, and a cosinor model was fitted to the median values per 15-minute interval. Cosinor mesor is not shown as normalization removes baseline level differences. Abbreviations: RMSE = root mean square of successive differences; SE = standard error;

| | Continuous group | Cyclic daytime group |
|---|---|---|
| **Core body temperature** | n=24 | n=18 |
| Amplitude (± SE) | 0.076 ± 0.009 | 0.084 ± 0.015 |
| Acrophase (± SE) | 20:32 ± 00:26 | 21:13 ± 00:41 |
| R2 | 0.28 | 0.14 |
| p-value of fit | <0.001 | <0.001 |
| **Heart rate** | n=22 | n=16 |
| Amplitude (± SE) | 0.56 ± 0.16 | 3.21 ± 0.20 |
| Acrophase (± SE) | 17:30 ± 01:05 | 17:46 ± 00:14 |
| R2 | 0.06 | 0.57 |
| p-value of fit | 0.003 | <0.001 |
| **Blood pressure** | n=29 | n=21 |
| Amplitude (± SE) | 0.75 ± 0.16 | 0.85 ± 0.24 |
| Acrophase (± SE) | 10:18 ± 00:48 | 06:25 ± 01:04 |
| R2 | 0.11 | 0.06 |
| p-value of fit | <0.001 | 0.003 |
| **HRV (RMSSD)** | n=22 | n=16 |
| Amplitude (± SE) | 0.119 ± 0.23 | 1.081 ± 0.292 |
| Acrophase (± SE) | 18:43 ± 07:21 | 11:38 ± 01:02 |
| R2 | 0.00 | 0.07 |
| p-value of fit | 0.875 | 0.001 |

**e-Table 5. Outcomes of cosinor and non-parametric analyses of 24-hour rhythms in vital signs (follow-up cohort, intention-to-treat analysis).**

Normally distributed continuous variables are presented as mean ± SD and compared between groups using a t-test, non-normally distributed variables are presented as median [IQR25–IQR75] and compared using the Mann–Whitney U test.

[a]: Group comparison was not performed because significant phase clustering (Rayleigh test) was observed in none groups or only one group.

Significant differences between groups are indicated as: p<0.05 (*), p<0.01 (**), and p<0.001 (***).

Abbreviations: IQR = Interquartile Range; RMSE = root mean square of successive differences; SD = Standard Deviation.

| | Continuous group | Cyclic daytime group | p-value |
|---|---|---|---|
| **Core body temperature** | n=27 | n=25 | |
| Cosinor amplitude (ºC), median [IQR] | 0.17 [0.10 - 0.24] | 0.20 [0.13 - 0.28] | 0.133 |
| Cosinor acrophase (hh:mm), circular mean ± circular SD (Rayleigh test p-value) | 18:24 ± 08:06 (p=0.745) | 00:25 ± 05:53 (p=0.099) | n/a[a] |
| Cosinor mesor (ºC), mean ± SD | 37.39 ± 0.64 | 37.26 ± 0.54 | 0.449 |
| **Heart rate** | n=24 | n=19 | |
| Cosinor amplitude (bpm), median [IQR] | 2.7 [1.4 - 3.8] | 3.9 [2.6 – 5.0] | 0.094 |
| Cosinor acrophase (hh:mm), circular mean ± circular SD (Rayleigh test p-value) | 19:49 ± 06:59 (p=0.433) | 18:18 ± 04:01 (p=0.001**) | n/a[a] |
| Cosinor mesor (bpm), mean ± SD | 84.2 ± 16.6 | 79.9 ± 14.7 | 0.378 |
| **Mean blood pressure** | n=32 | n=28 | |
| Cosinor amplitude (mmHg), median [IQR] | 3.4 [2.5 - 4.9] | 4.4 [2.7 - 6.4] | 0.251 |
| Cosinor acrophase (hh:mm), circular mean ± circular SD (Rayleigh test p-value) | 13:12 ± 07:14 (p=0.421) | 17:33 ± 10:12 (p=0.978) | n/a[a] |
| Cosinor mesor (mmHg), mean ± SD | 78.1 ± 10.7 | 80.0 ± 10.3 | 0.485 |
| **Heart rate variability (RMSSD)** | n=24 | n=19 | |
| Cosinor amplitude (ms), median [IQR] | 4.65 [3.41 - 6.76] | 6.37 [1.75 - 11.76] | 0.616 |
| Cosinor acrophase (hh:mm), circular mean ± circular SD (Rayleigh test p-value) | 08:31 ± 07:00 (p=0.443) | 13:36 ± 05:54 (p=0.179) | n/a[a] |
| Cosinor mesor (ms), median [IQR] | 15.21 [11.12 - 31.96] | 22.85 [11.00 - 57.88] | 0.372 |

**e-Table 6. Reasons for exclusion of melatonin profiles in melatonin midpoint analysis (per-protocol cohort).**

| | Continuous group (n=24) | Cyclic daytime group (n=17) |
|---|---|---|
| 6/7 or 7/7 samples below lower limit of detection | 4 (16.7%) | 3 (17.6%) |
| Continuous decline across sampling period | 3 (12.5%) | 2 (11.8%) |
| Continuous rise across sampling period | 0 (0%) | 1 (5.8%) |
| No variation (flat profile) | 1 (4.2%) | 0 (0%) |

**e-Table 7. Intention-to-treat analysis of melatonin.**

| | Continuous group (n=34) | Cyclic daytime group (n=28) | p-value |
|---|---|---|---|
| **Included in melatonin analysis** | n=27 | n=23 | |
| **Linear mixed effects model** | | | |
| Sampling time | | | <0.001 |
| Randomisation group | | | 0.782 |
| Randomisation group x Sampling time | | | 0.979 |
| **Day/night ratio, median [IQR]** | 0.80 [0.25-1.18] | 0.80 [0.25-1.18] | 0.960 |
| **Melatonin levels estimable** | 17 / 27 (63%) | 17 / 23 (74%) | |
| **Melatonin midpoints (hh:mm) (Rayleigh test p-value), circular mean ± circular SD** | 06:07 ± 00:41 (p<0.001) | 06:04 ± 00:52 (p<0.001) | 0.961 |

**e-Table 8. Infection type and sites (full cohort, intention-to-treat analysis).**

Only infections were considered that were detected after the start of the study intervention. a Counts and percentages per infection type do not sum to the total number of infections or 100%, as infections with secondary infections are counted in more than one category. b Counts and percentages per infection type do not sum to the total number of infections or 100%, as pathogens detected at multiple sites are counted in more than one category.

| | Continuous group (n=47) | Cyclic daytime group (n=38) |
|---|---|---|
| **Total number of patients with infections, n (%)** | 25 (53.2%) | 20 (52.6%) |
| **Type of infection, n (%)[a]** | | |
| Bacterial | 28 (75.7%) | 24 (85.7%) |
| Viral | 5 (13.5%) | 2 (7.1%) |
| Fungal | 6 (16.2%) | 3 (10.7%) |
| **Infection sites, n (%)[b]** | | |
| Respiratory | 20 (54.1%) | 14 (50.0%) |
| Wound/surgical site | 0 (0.0%) | 1 (3.6%) |
| Urinary tract | 1 (2.7%) | 0 (0.0%) |
| Skin and soft tissue | 1 (2.7%) | 3 (10.7%) |
| Central nervous system | 0 (0.0%) | 1 (3.6%) |
| Catheter-related blood stream infections | 2 (5.4%) | 3 (10.7%) |
| Not-catheter-related blood stream infection | 13 (35.1%) | 7 (25.0%) |
| Device-related infections | 4 (10.8%) | 2 (7.1%) |
| Intra-abdominal | 6 (16.2%) | 0 (0.0%) |

### Supplementary Figures

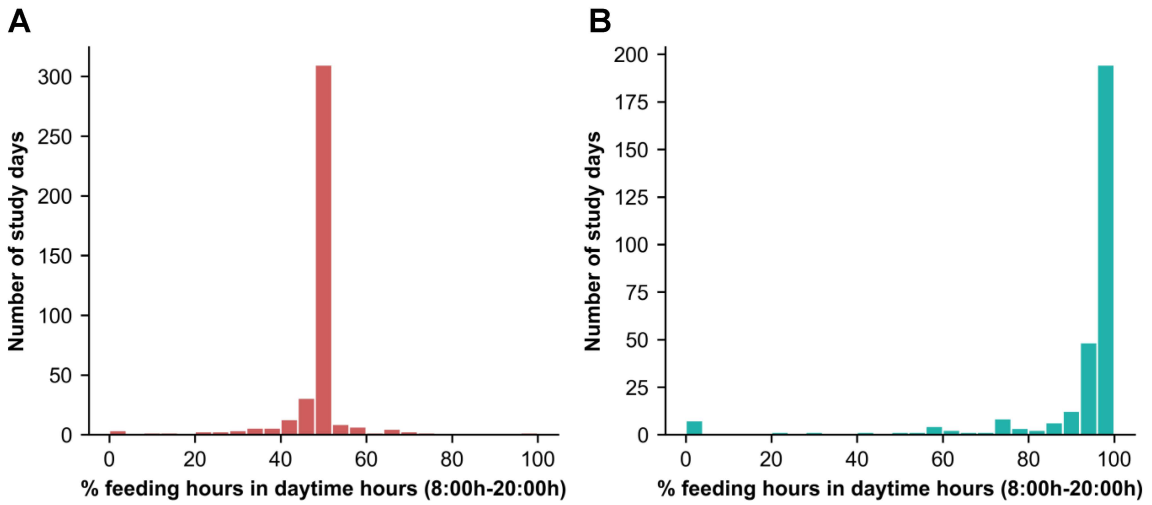

**e-Figure 1. Timing of nutrition and volume rates per study day.** **(A-B)** Feeding hours in daytime hours (8:00h-20:00h) across all study days in the **(A)** continuous group and **(B)** cyclic daytime group.

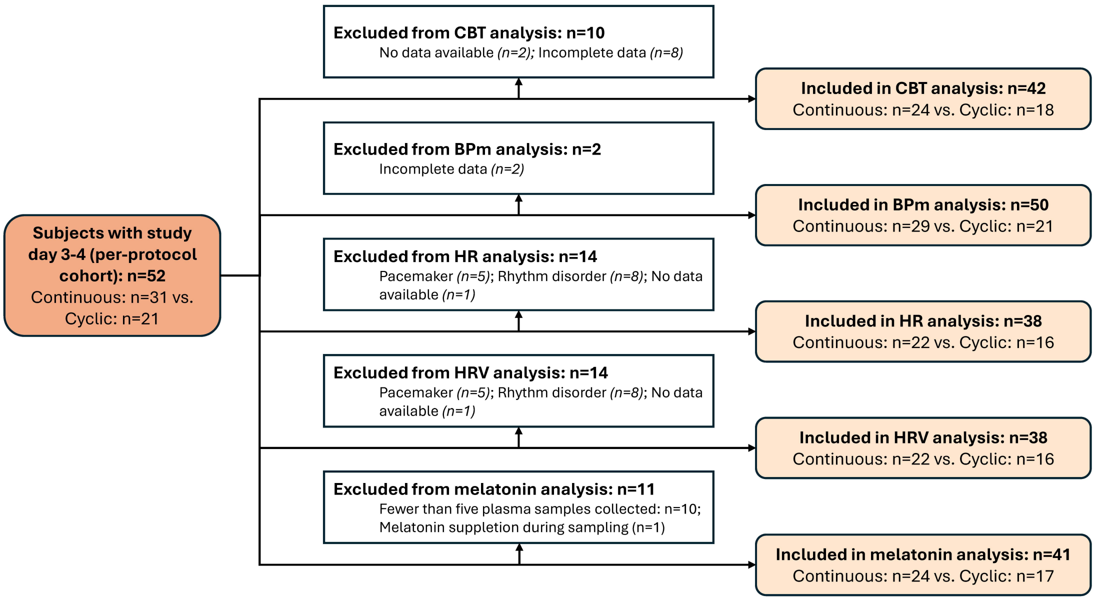

**e-Figure 2. Availability of vital sign and melatonin data (per-protocol cohort).**

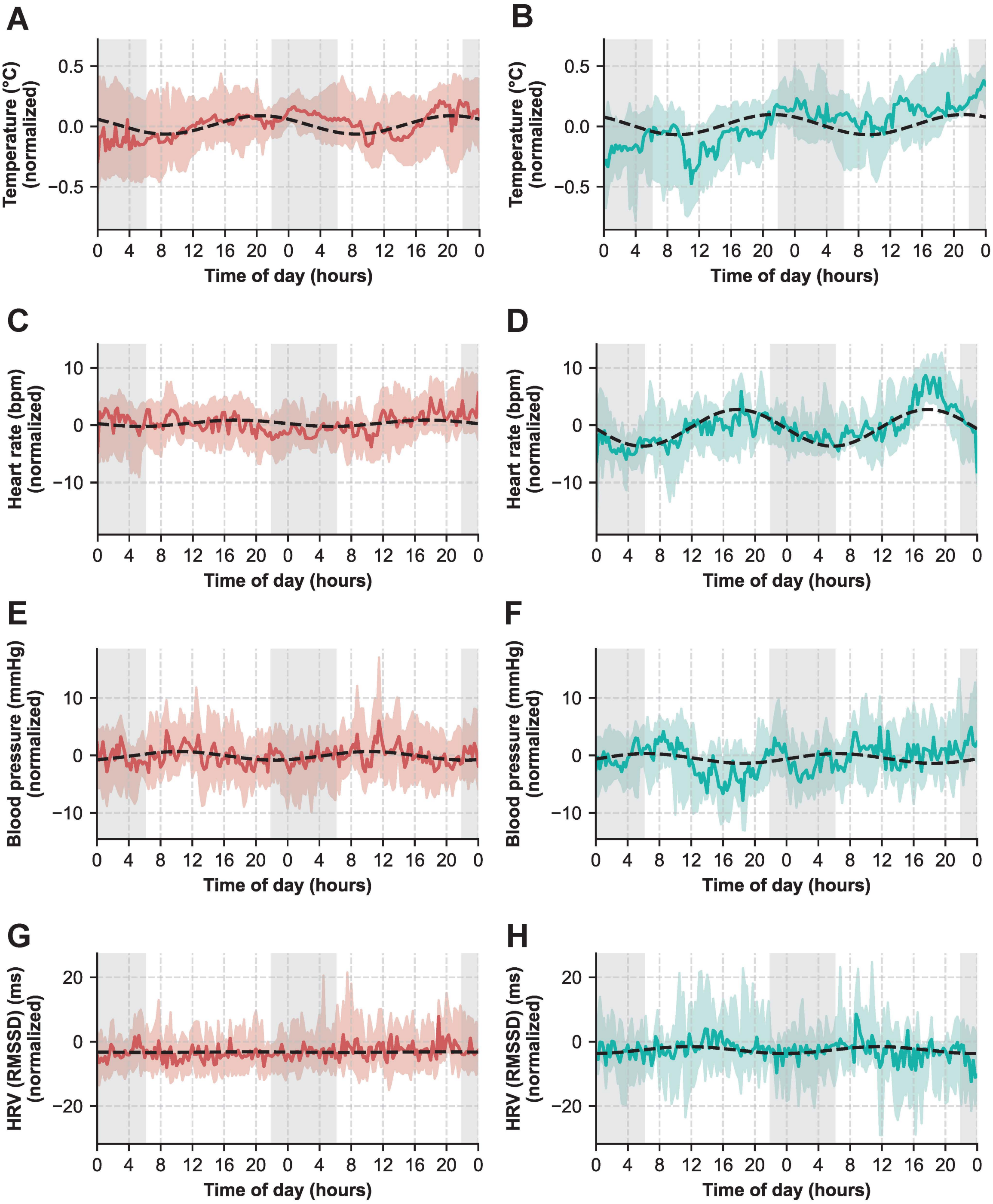

**e-Figure 3. Group-level visualization of 24-hour rhythms in vital signs across feeding groups** (follow-up cohort, per-protocol analysis). (A-B) Core body temperature for (A) continuous group (n=24) and (B) cyclic daytime group (n=18). (C-D) Heart rate for (C) continuous group (n=22) and (D) cyclic daytime group (n=16). (E-F) Mean arterial blood pressure for (E) continuous group (n=29) and (F) cyclic daytime group (n=21). (G-H) Heart rate variability (RMSSD) for (G) continuous group (n=22) and (H) cyclic daytime group (n=16). Data was normalized by mean subtraction per individual and is shown as median and interquartile range across each 15-minute interval. The black dashed line represents a cosinor fit on the aggregated time series with median values across groups. Cosinor parameters for each cosinor fit are provided in e-Table 3. Gray shaded areas represent nighttime hours (22:00h-6:00h).

### References

1. Van Rossum GDJ, Fred L. Python reference manual: Centrum voor Wiskunde en Informatica Amsterdam; 1995.
2. Pan J, Tompkins WJ. A real-time QRS detection algorithm. *IEEE Trans Biomed Eng* 1985; **32**(3): 230-6.
3. Bota P, Silva R, Carreiras C, Fred A, da Silva HP. BioSPPy: A Python toolbox for physiological signal processing. *Softwarex* 2024; **26**.
4. Rebergen DJ, Nagaraj SB, Rosenthal ES, Bianchi MT, van Putten M, Westover MB. ADARRI: a novel method to detect spurious R-peaks in the electrocardiogram for heart rate variability analysis in the intensive care unit. *J Clin Monit Comput* 2018; **32**(1): 53-61.
5. Gomes PMP, Placido da Silva, Hugo. pyHRV: Development and Evaluation of an Open-Source Python Toolbox for Heart Rate Variability (HRV). International Conference on Electrical, Electronic and Computing Engineering (IcETRAN)At: Serbia; 2019. p. 822–8.
6. van Faassen M, Bischoff R, Kema IP. Relationship between plasma and salivary melatonin and cortisol investigated by LC-MS/MS. *Clin Chem Lab Med* 2017; **55**(9): 1340-8.
7. Brown EN, Czeisler CA. The statistical analysis of circadian phase and amplitude in constant-routine core-temperature data. *J Biol Rhythms* 1992; **7**(3): 177-202.
8. Bowman C, Huang Y, Walch OJ, et al. A method for characterizing daily physiology from widely used wearables. *Cell Rep Methods* 2021; **1**(4).
9. Pinheiro JC, Bates D. nlme: Linear and Nonlinear Mixed Effects Models. 2025.
10. Benloucif S, Burgess HJ, Klerman EB, et al. Measuring melatonin in humans. *J Clin Sleep Med* 2008; **4**(1): 66-9.
11. Badawi O, Yeung SY, Rosenfeld BA. Evaluation of glycemic control metrics for intensive care unit populations. *Am J Med Qual* 2009; **24**(4): 310-20.
12. Khan BA, Perkins AJ, Gao S, et al. The Confusion Assessment Method for the ICU-7 Delirium Severity Scale: A Novel Delirium Severity Instrument for Use in the ICU. *Crit Care Med* 2017; **45**(5): 851-7.
